# Toward personalized skin cancer care: multiple skin cancer development in five cohorts

**DOI:** 10.1101/2024.05.06.24306947

**Authors:** Lee Wheless, Kai-Ping Liao, Siwei Zheng, Yao Li, Lydia Yao, Yaomin Xu, Christopher Madden, Jacqueline Ike, Isabelle T Smith, Dominique Mosley, Sarah Grossarth, Rebecca I Hartman, Otis Wilson, Adriana Hung, Mackenzie R Wehner

## Abstract

**Importance:** Many patients will develop more than one skin cancer, however most research to date has examined only case status.

**Objective:** Describe the frequency and timing of the treatment of multiple skin cancers in individual patients over time

**Design:** Longitudinal claims and electronic health record-based cohort study

**Setting:** Vanderbilt University Medical Center database called the Synthetic Derivative, VA, Medicare, Optum Clinformatics® Data Mart Database, IBM Marketscan

**Participants:** All patients with a Current Procedural Terminology code for the surgical management of a skin cancer in each of five cohorts.

**Exposures:** None.

**Main Outcomes and Measures:** The number of CPT codes for skin cancer treatment in each individual occurring on the same day as an ICD code for skin cancer over time

**Results:** Our cohort included 5,508,374 patients and 13,102,123 total skin cancers treated.

**Conclusions and Relevance:** Nearly half of patients treated for skin cancer were treated for more than one skin cancer. Patients who have not developed a second skin cancer by 2 years after the first are unlikely to develop multiple skin cancers within the following 5 years. Better data formatting will allow for improved granularity in identifying individuals at high risk for multiple skin cancers and those unlikely to benefit from continued annual surveillance. Resource planning should take into account not just the number of skin cancer cases, but the individual burden of disease.

**Key points:** Question: How many skin cancer patients are treated for more than one skin cancer and how soon after the first skin cancer do they occur?

Findings: 43% of patients were treated for more than one skin cancer, the majority of which occurred within two years after the initial skin cancer. Just 3% of patients were treated for 10 or more skin cancers, but these patients accounted for 22% of all of the skin cancer treatments in the cohort Meaning: Nearly half of all skin cancer patients were treated for multiple skin cancers, while those without a second skin cancer after two years were less likely to be treated for a subsequent skin cancer within the next five years.

## INTRODUCTION

Keratinocyte carcinomas are excluded from cancer registries in the United States, and often only the first cancer is registered in countries that do include them^1–4^. As a result, high quality information regarding multiplicity of skin cancer can be difficult to obtain. Several studies in small populations or high-risk groups have shown that the risk of developing a second keratinocyte carcinoma is up to 60% within 10 years after the initial skin cancer, indicating that multiple skin cancers are a common problem^5–9^. Prior data suggests that individuals with multiple skin cancers are more likely to develop metastases, harbor mutations that predispose to other malignancies, or experience skin cancer-related death^10,11^. The number of skin cancers a patient will develop thus has clinical importance beyond simply the burden of treating the skin cancers themselves. Moreover, understanding who is and is not likely to develop multiple skin cancers could aid in resource allocation and planning by identifying those who would benefit most from more aggressive screening and chemoprevention, as well as adjusting screening intervals to improve access to dermatologic care. We conducted this study to describe the frequency and timing of multiple skin cancers and provide individual-level counts for multiple skin cancer development.

## METHODS

### Cohorts

This project was approved by the institutional review boards (IRB) of Vanderbilt University Medical Center (VUMC) (non-human subjects designation, IRB# 200335), Tennessee Valley Health System Veterans Administration Medical Center (exempt status, IRB# 1657284), and MD Anderson Cancer Center (IRB# 2019-0966). We used five large electronic health record (EHR) cohorts: VUMC’s de-identified research database called the Synthetic Derivative (VUMC, 1/1989-9/2023), the Veteran’s Administration Informatics and Computing Infrastructure (VINCI, 10/1999-2/2024)^12^, the Centers for Medicare Services dataset of 4,999,999 fee-for-service beneficiaries with Part D coverage (Medicare; 2009-2018), Optum’s de-identified Clinformatics® Data Mart Database (Optum; 1/2007 - 6/2022), and the Merative^TM^ MarketScan® Research Database (MarketScan; 1998-2021). From each cohort we collected information on demographics including self-reported race, dates of service, diagnostic and procedural codes, and medications. The main outcome was the development of skin cancer as measured by the entry of a Current Procedural Terminology (CPT) code for the treatment of skin cancer entered on the same day as an International Classification of Disease (ICD)-9 or -10 code for skin cancer (Supplemental Table 1)^13^. The date of entry for each CPT code was used to keep a running tally of the number of skin cancers per individual over time. The earliest ICD code date for any condition was the time of patient entry into each cohort, and the latest ICD code date for any condition or date of death where applicable was the end of follow-up for each patient. All time intervals were calculated relative to the date of the first skin cancer code per patient.

Each cohort contained different data fields. In preliminary analyses across cohorts, many of the skin cancer ICD codes were not specific for a type of skin cancer (e.g. ICD-10 C44.90 “Unspecified malignant neoplasm of the skin, unspecified”) and many patients had more than one skin cancer ICD code on the same date, so no further typing was attempted in this project^14^. We excluded patients with >150 separate CPT codes for skin cancer treatment and those with >15 CPT codes in a single day, as these were felt to be either data errors or potentially identifiable data if correct. Patients could have more than one skin cancer treatment on the same day, as is common clinical practice. Each skin cancer treatment CPT code was treated as an individual skin cancer in all analyses.

Not all cohorts had long follow-up or complete records. VUMC contains data from an open health care system in which not all data are captured by the VUMC EHR, Medicare does not contain data prior to enrollment, usually at age 65 or older, and MarketScan and Optum contain numerous intervals of patients coming into and out of the database. Therefore, we restricted analyses including follow-up intervals to VINCI to ensure sufficient data capture. VINCI began collecting data 10/1/1999, although records for skin cancer treatment exist from as early as 1994. All instances of skin cancer treatment were included in our counts as the goal was to present the most complete data possible.

### Statistical Analysis

Descriptive statistics for each cohort were calculated, with any differences in means and proportions between groups compared using t-tests and χ^2^ tests, as appropriate. To compare mean number of skin cancers by age at first skin cancer between cohorts, we used ANOVA. To evaluate whether patients with more years of follow-up had greater numbers of skin cancers in VINCI, we used Pearson’s correlation coefficient to measure linear association. Mean cumulative function plots were constructed to estimate the mean number of skin cancers per patient over time using the reda package in R or proc reliability in SAS^15^. All analyses were conducted in R v4.2.2 or SAS version 9.4. Analyses were conducted between 10/2023 and 2/2024.

## RESULTS

Overall, there were 5,508,374 total patients with skin cancer included in the study, with 13,102,123 total treated skin cancers. The mean age at first skin cancer was between 63 and 76 (overall mean 66.1 years, standard deviation 10.1 years). There was a slight male predominance overall when excluding the VA population, which was almost exclusively male, and greater than 90% of patients reported their race as White (Table 1). Each database covered a period of 10 to 36 years. There were 3,152,904 (57.2%) individuals who were treated for a single skin cancer during their follow-up intervals, while 2,355,470 (42.8%) were treated for more than one skin cancer, and 165,745 (3.0%) were treated for at least ten skin cancers. Those 3.0% treated for at least ten skin cancers contributed 2,855,555, or 21.8% of all skin cancers. Those who ultimately were treated for 10 or more skin cancers on average were treated for their first skin cancer more than 2 years earlier in age than those who were treated for fewer skin cancers (68.0 vs 70.6, p < 0.01). There was significant variability across cohorts in the mean number of skin cancers treated based on age at first skin cancer (p < 0.01, Table 2). In general, patients whose first skin cancer was treated before age 30 developed fewer than those whose first skin cancer was treated after age 50. Among those who did develop a second skin cancer, the median time to second skin cancer was one year or less for nearly all age groups across all cohorts (Table 3).

**Table 1.** Characteristics of individuals in each cohort.

|  | VUMC<sup>1</sup> (n = 41,527) | VINCI<sup>2</sup> (n = 607,814) | Medicare (n=819,266) | Optum (n=1,951,065) | MarketScan (n=2,088,702) |
| --- | --- | --- | --- | --- | --- |
| Mean age at first skin cancer (SD<sup>3</sup>) | 64.1 (13.6) 65.4 | 70.6 (11.1) | 76.3 (8.3) | 68.5 (12.7) | 53.6 (8.4) |
| Sex at birth | Male: 23,393 (56.3%)<br>Female: 18,130 (43.7%)<br>Other: 4 (<0.1%) | Male: 592,098(97.4%)<br>Female: 15,716(2.6%) | Male: 396010 (48.3%)<br>Female: 423256 (51.7%) | Male: 1,067,224 (54.7%)<br>Female: 883,681 (45.3%)<br>Other: 160 (0.01%) | Male: 1,075,384 (51.5%)<br>Female: 1,013,318 (48.5%) |
| Race and ethnicity | NHW<sup>4</sup>: 37,233 (89.7%)<br>HW<sup>5</sup>: 18 (<0.1%)<br>B<sup>6</sup>: 135 (<0.1%)<br>A<sup>7</sup>: 39 (<0.1%)<br>O<sup>8</sup>: 4,102 (9.9%) | NHW: 541,587 (89.1%)<br>HW: 9,118 (1.5%)<br>B: 5,698 (0.9%)<br>A: 477 (0.1%)<br>O: 50,934 (8.4%) | NHW: 2989301 (97.6%)<br>H: 29090 (1.0%)<br>B: 3366 (0.1%)<br>A: 7087 (0.2%)<br>O: 32110 (1.1%)<br>Unknown: 8011 (1.0%) | W: 1,680,231 (86.1%)<br>B: 80,078 (4.1%)<br>H: 66,444 (3.4%)<br>A: 13,864 (0.7%)<br>Unknown: 110,448 (5.7%) | N/A |
| Earliest date of first skin cancer treatment code | 8/8/1997 | 12/20/1994 | 1/1/2009 | 1/1/2007 | 1/2/1998 |
| Latest date of skin cancer treatment | 8/29/2023 | 2/3/2024 | 12/31/2018 | 6/30/2022 | 12/31/21 |
| Mean years of follow-up after first skin cancer (SD<sup>3</sup>) | 2.7 (4.3) | 9.3 (5.9) | 4.58 (2.89) | 2.33 (2.61) | 2.88 (3.12) |
| Mean years of follow-up before first skin cancer | 0.4 (1.6) | 6.5 (5.5) | 4.11 (2.89) | 4.36 (3.88) | 2.90 (3.32) |
| (SD<sup>3</sup>) |  |  |  |  |  |
| Number of skin cancers | Total: 93,824 | Total: 1,515,553 | Total: 3,064,070 | Total: 4,758,758 | Total: 3,669,918 |
|  | 1: n = 25,966 (62.5%) | 1: n = 316,327 (52.0%) | 1: n = 329,068 (40.2%) | 1: n = 1,079,571 (55.3%) | 1: n = 1,401,972 (67.1%) |
|  | 2-3: n = 10,126 (24.4%) | 2-3: n = 185,159 (30.5%) | 2-3: n = 256,494 (31.3%) | 2-3: n = 557,889 (28.6%) | 2-3: n = 519,608 (24.9%) |
|  | 4-9: n = 4,183 (10.1%) | 4-9: n = 87,244 (14.4%) | 4-9: n = 170,569 (20.8%) | 4-9: n = 252,318 (12.9%) | 4-9: n = 146,135 (7.0%) |
|  | 10+: n = 1,252 (3.0%) | 10+: n = 19,084 (3.1%) | 10+: n = 63,135 (7.7%) | 10+: n = 61,287 (3.1%) | 10+: n = 20,987 (1.0%) |
| 1- Vanderbilt University Medical Center Synthetic Derivative Cohort<br>2- Veterans Affairs Informatics and Computing Infrastructure Cohort<br>3- Standard deviation<br>4- Self-reported non-Hispanic White<br>5- Self-reported Hispanic White<br>6- Self-reported Black<br>7- Self-reported Asian<br>8- Self-reported other race and ethnicity |  |  |  |  |  |

**Table 2.** Mean number of skin cancers by age at first skin cancer.

|  | VUMC<sup>1</sup> (n = 41,527) | VINCI<sup>2</sup> (n = 576,460) | Optum (n=1,951,065) | MarketScan (n=2,088,702) |
| --- | --- | --- | --- | --- |
| Mean number of skin cancers (SD<sup>3</sup>) | 20-29: 1.8 (5.6) | 20-29: 1.3 (1.1) | 20-29: 1.3 (1.2) | 20-29: 1.4 (1.5) |
|  | 30-39: 1.8 (3.9) | 30-39: 1.7 (2.8) | 30-39: 1.5 (1.8) | 30-39: 1.6 (1.9) |
|  | 40-49: 2.1 (3.7) | 40-49: 2.3 (3.7) | 40-49: 1.8 (2.5) | 40-49: 1.8 (2.5) |
|  | 50-59: 2.3 (3.8) | 50-59: 2.8 (4.2) | 50-59: 2.0 (2.8) | 50-59: 1.9 (2.3) |
|  | 60-69: 2.5 (3.9) | 60-69: 2.7 (3.4) | 60-69: 2.4 (3.5) | 60-69: 1.6 (1.4) |
|  | 70+: 2.2 (2.9) | 70+: 2.4 (2.8) | 70+: 2.7 (3.8) | 70+: 2.1 (2.3) |
| 1- Vanderbilt University Medical Center Synthetic Derivative Cohort |  |  |  |  |
| 2- Veterans Affairs Informatics and Computing Infrastructure Cohort |  |  |  |  |
| 3- Standard deviation |  |  |  |  |

**Table 3.**
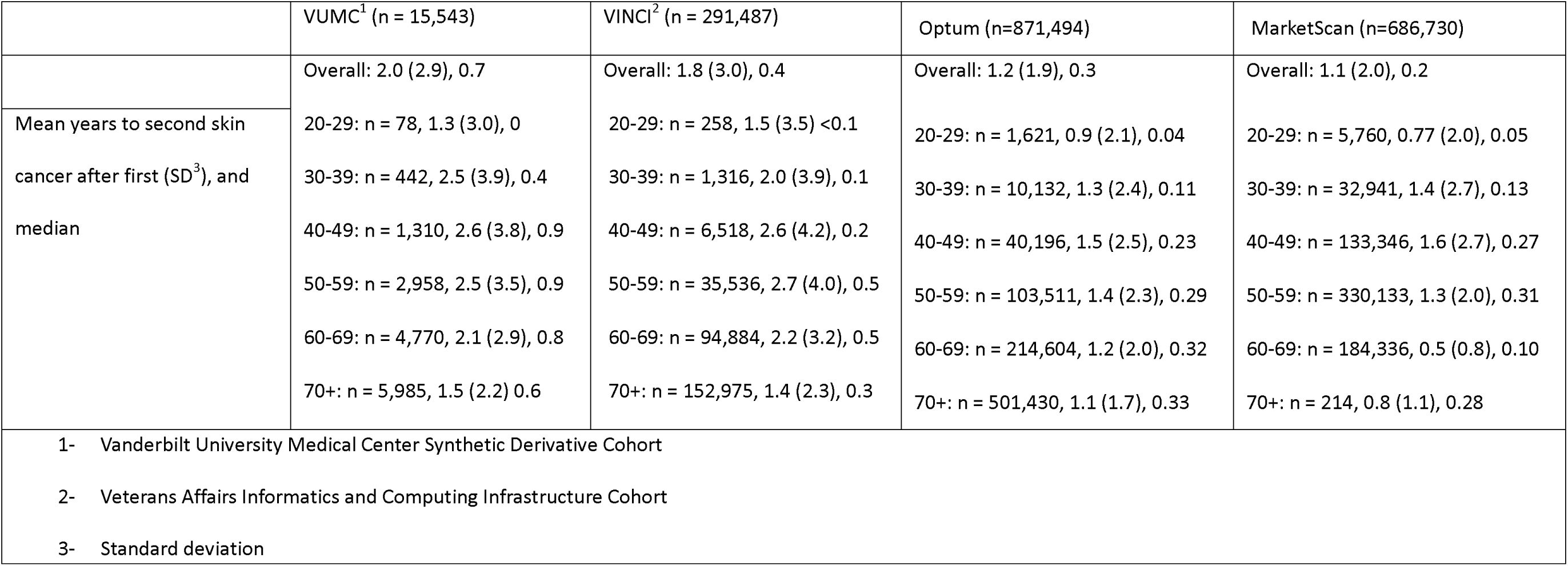
Time to second skin cancer by age at first skin cancer.

|  | VUMC<sup>1</sup> (n = 15,543) | VINCI<sup>2</sup> (n = 291,487) | Optum (n=871,494) | MarketScan (n=686,730) |
| --- | --- | --- | --- | --- |
|  | Overall: 2.0 (2.9), 0.7 | Overall: 1.8 (3.0), 0.4 | Overall: 1.2 (1.9), 0.3 | Overall: 1.1 (2.0), 0.2 |
| Mean years to second skin cancer after first (SD<sup>3</sup>), and median | 20-29: n = 78, 1.3 (3.0), 0<br>30-39: n = 442, 2.5 (3.9), 0.4<br>40-49: n = 1,310, 2.6 (3.8), 0.9<br>50-59: n = 2,958, 2.5 (3.5), 0.9<br>60-69: n = 4,770, 2.1 (2.9), 0.8<br>70+: n = 5,985, 1.5 (2.2) 0.6 | 20-29: n = 258, 1.5 (3.5) <0.1<br>30-39: n = 1,316, 2.0 (3.9), 0.1<br>40-49: n = 6,518, 2.6 (4.2), 0.2<br>50-59: n = 35,536, 2.7 (4.0), 0.5<br>60-69: n = 94,884, 2.2 (3.2), 0.5<br>70+: n = 152,975, 1.4 (2.3), 0.3 | 20-29: n = 1,621, 0.9 (2.1), 0.04<br>30-39: n = 10,132, 1.3 (2.4), 0.11<br>40-49: n = 40,196, 1.5 (2.5), 0.23<br>50-59: n = 103,511, 1.4 (2.3), 0.29<br>60-69: n = 214,604, 1.2 (2.0), 0.32<br>70+: n = 501,430, 1.1 (1.7), 0.33 | 20-29: n = 5,760, 0.77 (2.0), 0.05<br>30-39: n = 32,941, 1.4 (2.7), 0.13<br>40-49: n = 133,346, 1.6 (2.7), 0.27<br>50-59: n = 330,133, 1.3 (2.0), 0.31<br>60-69: n = 184,336, 0.5 (0.8), 0.10<br>70+: n = 214, 0.8 (1.1), 0.28 |
| 1- Vanderbilt University Medical Center Synthetic Derivative Cohort<br>2- Veterans Affairs Informatics and Computing Infrastructure Cohort<br>3- Standard deviation |  |  |  |  |

### VINCI-specific analyses

In the VINCI cohort, there was a linear increase in the mean number of skin cancers per patient over time (r =0.28, p < 0.01), with patients on average having been treated for 1.4 more skin cancers within two years after the initial skin cancer, and 1.9 within five years, similar to other cohorts with less complete data capture (Supplemental Figures 1-5). These rates differed based on age at first skin cancer, with faster rates of accumulation occurring among those developing their first skin cancer at older ages (Figure 1, p < 0.1). The mean rate at which patients developed skin cancers was initially elevated in the two years following the first skin cancer, then remained stable among patients who were treated for fewer than ten skin cancers, while patients who were treated for ten or more skin cancers had rates persistently 2-3 times those for other groups across a 20-year period (Supplemental Figure 6). Of patients who were treated for a second skin cancer, 71% of these occurred within 2 years after the first. Of patients who made it to 2, 2.5, and 3 years after the initial skin cancer without a second, 26.0%, 24.6% and 23.3%, respectively, developed an additional skin cancer in subsequent 5 years. Of the patients who made it to 2 years after the initial skin cancers without developing a second, only 1.1% (3,484/322,571) ultimately developed 10 or more, with most who were treated for additional skin cancers (56,255/83,902, 67.0%) having only 1 or 2 more. To explore individual-level incidence, we constructed spaghetti plots of cumulative skin cancers treated over time per patient (Figure 2). Only those with >50 skin cancer treatments are shown to allow for discrimination of individual lines. Patients who developed a high number of skin cancers tended to have a near-linear increase skin cancer treatments, while several developed many in a short period.

**Figure 1.**
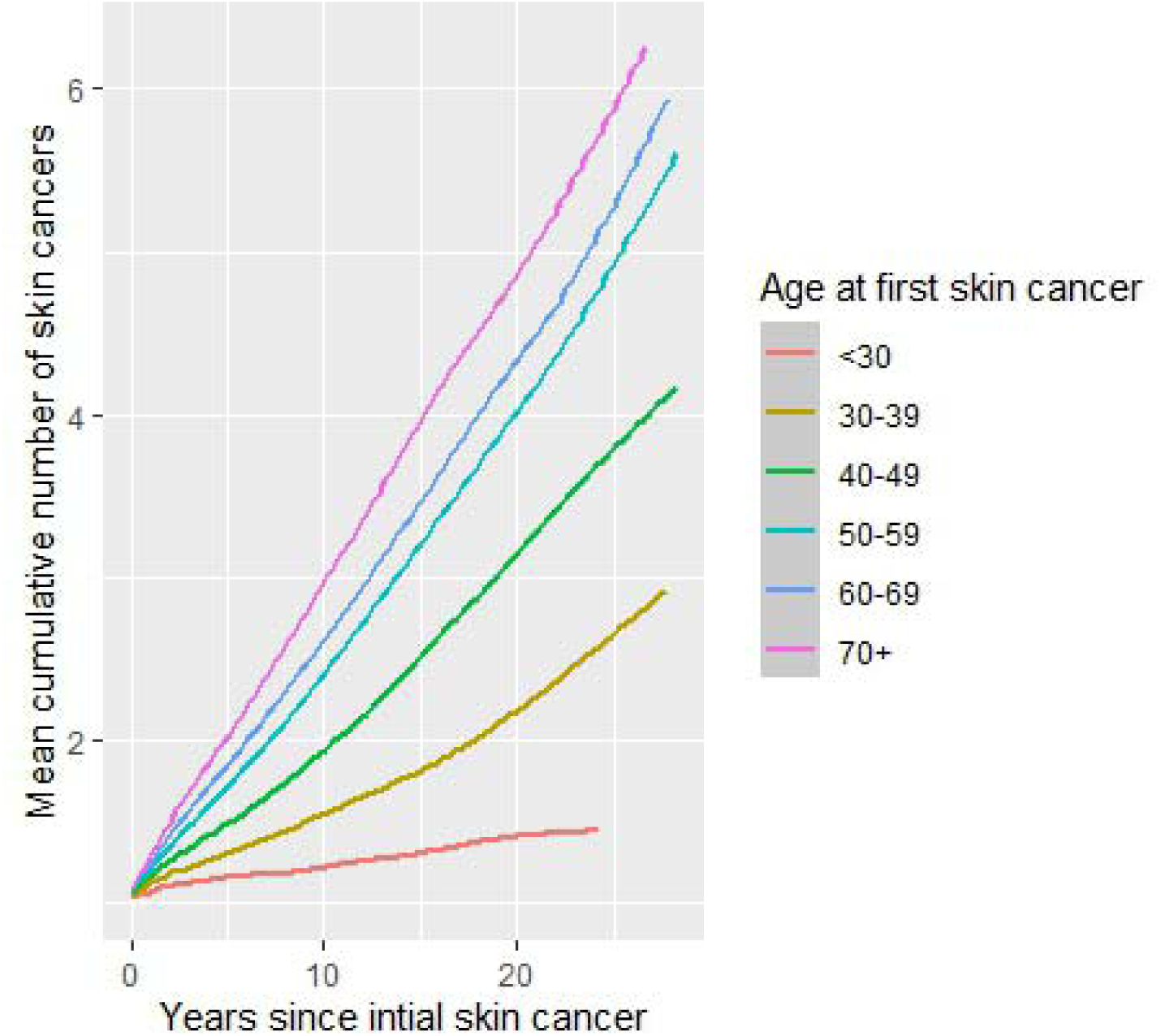
Mean cumulative function for the total number of skin cancers over time among patients in the overall VINCI cohort stratified by patient age at first skin cancer.

**Figure 2.**
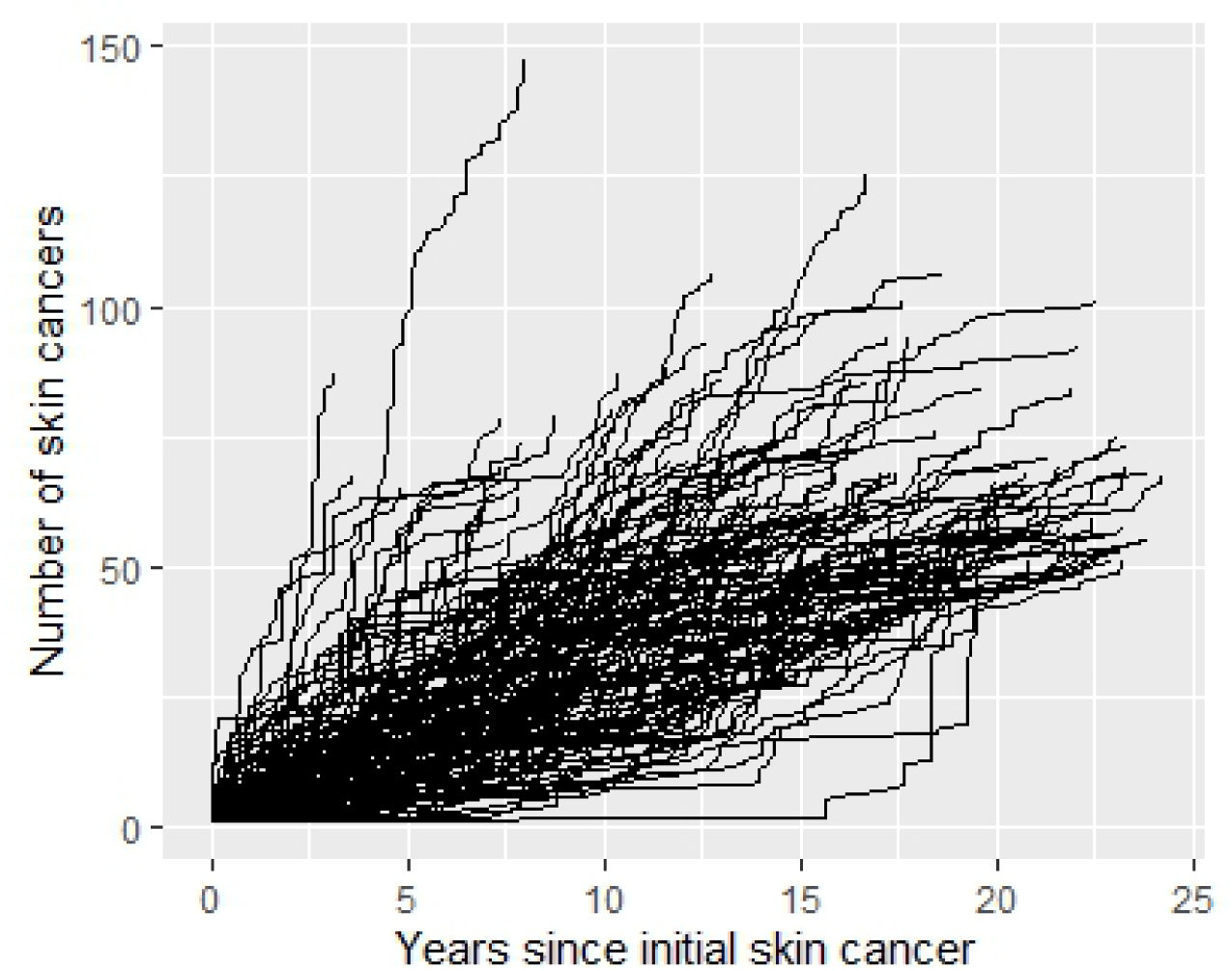
Individual-level cumulative incidence of skin cancers over time among patients in the VINCI cohort with 50 or more skin cancers treated.

## DISCUSSION

In our cohort study of more than 5.5 million patients treated for skin cancer in the United States over greater than two decades, we observed that 43% of all patients treated for one skin cancer will be treated for another and that greater than 4% will be treated for ten or more skin cancers. This very small subset with ten or more contributed one quarter of all skin cancers treated in the study and represent a population that demands early identification. There was consistency across cohorts in both the proportion of having more than one skin cancer surgery and the proportion of having ten or more. In each cohort, the median time to second cancer was less than one year for those who developed a subsequent skin cancer. This interval is consistent with the National Comprehensive Cancer Center guidelines of screening patients with a skin cancer every 3-12 months^16–19^.

Clinically, dermatologists can anecdotally appreciate how common it is for a patient to develop multiple skin cancers, often having more than one biopsied during the same appointment. Early and effective preventive intervention among those with the greatest skin cancer burden has the potential for the greatest benefit. Future studies will be needed to identify patients who will not just become skin cancer cases, but who will develop high numbers of skin cancer cases. Just as important as identifying high risk patients is identifying low risk patients who are unlikely to benefit from annual surveillance^16–19^. To evaluate the possibility of identifying low risk patients, we investigated skin cancer rates after the initial cancer in the VINCI cohort. We observed an initial increase in the rate of skin cancer that decayed to a steady state over two years (Figure 2). We therefore examined patients who made it beyond this initial two-year period of increased skin cancer risk without a subsequent one to determine if their risk of subsequent skin cancers was sufficiently low to widen their follow-up interval. One in four patients who had a 2-year gap after the first skin cancer was later treated for a subsequent skin cancer, a proportion still too high to make clinical recommendations regarding reduced intensity surveillance. However, future studies to better identify patients at lowest risk could improve our ability to personalize and de-escalate follow-up intervals. Patients with large, aggressive, or metastatic skin cancers should continue skin surveillance at regular intervals as our cohorts were not equipped to address these special populations.

Patients with large numbers of skin cancers are clinically important for multiple reasons. First, these patients experience a greater burden of disease than patients with only one or two, with increased morbidity, risk of metastasis and mortality, and financial impacts. Next, patients with multiple skin cancers are more likely to harbor mutations in DNA repair genes, or to develop internal cancers^11^. Lastly, a study of solid organ transplant recipients showed that patients with 10 or more skin cancers contributed nearly half of all skin cancer-related deaths in this population^10^. Without well-curated, validated data on skin cancer, there has been limited ability to identify these patients for more aggressive prevention, or assess preventive measures in pragmatic trials^20–22^. Data from Europe support prior estimates in the US showing that deaths from keratinocyte carcinomas now outnumber those from melanoma^23,24^. Our results are a call to action for renewed focus on these most common cancers in the United States.

Handling skin cancer as a binary outcome is a hindrance to precision medicine and creates missed opportunities for clinical intervention. Moreover, resource allocation and reimbursement should account for the fact that nearly 1 in 2 patients treated for skin cancer will be treated for a second one, with a median time of less than one year after the first. Finally, in the era of large language models, automated systems for high-throughput data formatting are increasingly being used, facilitating rapid extraction of both prospective and historical data. Database structures and data collection should be modified to allow for skin cancer multiplicity and increased granularity.

Our study had multiple limitations. First, we used same-day ICD and CPT codes to count skin cancers. We have previously shown that using a CPT code for skin cancer treatment has good correlation with the number of histologically-verified skin cancers. Next, we had minimal inclusion and exclusion criteria with widely variable follow-up periods and ages of coverage in the datasets. As a result, we are likely underestimating both the frequency and counts for skin cancers in this study. Instead, we sought to capture as many patients as possible who were treated for multiple skin cancers. Patients with more skin cancers treated had more years of follow-up, suggesting that there might have been a follow-up bias in our results for how many skin cancers a patient would be treated. In VINCI, the median follow-up for those with a single skin cancer was 5.6 years, a time by which greater than 90% percent of patients with at least two skin cancers had already been treated for a second, which indicates these patients would most likely have developed additional skin cancers during this interval if they were to develop more than one. Moreover, we saw persistently elevated rates of skin cancer among those with ten or more at every time point across a 20-year period (Figure 2), which suggests that differences in follow-up time by the number of skin cancers per patient were unlikely to explain the overall patterns observed. Our methodology would also capture treatment of recurrences and in transit metastases, which could inflate the number of skin cancers per patient. Given that these are both rare events, they are unlikely to impact the overall conclusions. While it is certain that some individuals are represented in multiple cohorts, such as VA patients seen at the Tennessee Valley VA Medical Center referred to VUMC for Mohs surgery, the de-identified nature of these datasets precludes any ability to assess and correct for duplication. We presented data from the cohorts separately, and the finest granularity analyses were conducted in only one cohort to avoid counting individuals twice. Finally, none of the cohorts used in this study are population-based and are therefore inadequate for drawing any conclusions regarding exact incidence or prevalence of multiple skin cancers. However, our findings are consistent with those from smaller population-based and studies of organ transplant recipients that also showed the development of multiple skin cancers is a common problem^10,25^.

## Conclusions

In this large consortium cohort of greater than 5 million patients with skin cancer, we observed that nearly half of all patients treated for skin cancer will be treated for at least one more. A subset of only 3% of patients contributed 22% of all skin cancers treated in this study. There is a critical need for improved data collection on skin cancers. Future efforts should be twofold: 1) early identification of these highest risk patients, and 2) identification of patients unlikely to benefit from routine annual surveillance who can safely be monitored at less frequent intervals. These goals require the development of improved systems for the processing and capture of skin cancer data.

## Data Availability

Data for each cohort are access-protected and cannot be shared externally in accordance with the authors' data use agreements

## Abbreviations

CPT: current procedural terminology
EHR: electronic health record
ICD: international classification of disease
IRB: Institutional Review Board
VINCI: Veteran’s Administration Informatics and Computing Infrastructure
VUMC: Vanderbilt University Medical Center

## Conflict of interest

The authors have no conflicts to declare

Study approved as VUMC IRB #200335 exempt non-human subjects research, VA IRB 1657284 exempt research, and MDA IRB 2019-0966 approved.

## Funding and Acknowledgements

*Dr. Wheless is supported by VA CSR&D grant IK2 CX002452. Dr. Hartman is supported by VA CSR&D grant IK2 CX002531. Dr. Wehner is supported by NCI grant K08CA263298. This research was supported by Cancer Prevention and Research Institute of Texas RR190078 (MRW) and a grant from the University of Texas Rising STARs program (MRW), as well as, in part, by Cancer Center Support Grant P30 CA016672 to MD Anderson Cancer Center. MRW is a Cancer Prevention and Research Institute of Texas Scholar in Cancer Research. The funder had no role in the design and conduct of the study; collection, management, analysis, and interpretation of the data; preparation, review, or approval of the manuscript; and decision to submit the manuscript for publication. The project described was supported by CTSA award No.* UL1TR000445 *from the National Center for Advancing Translational Sciences. Its contents are solely the responsibility of the authors and do not necessarily represent official views of the National Center for Advancing Translational Sciences, the National Institutes of Health, the Department of Veteran Affairs, or the United States Government*.

## Notes

### Competing Interest Statement

The authors have declared no competing interest.

### Author Declarations

This project was approved by the institutional review boards (IRB) of Vanderbilt University Medical Center (VUMC) (non-human subjects designation, IRB# 200335), Tennessee Valley Health System Veterans Administration Medical Center (exempt status, IRB# 1657284), and MD Anderson Cancer Center (IRB# 2019-0966).

