## Supplemental Figures for "Toward personalized skin cancer care: multiple skin cancer development in five cohorts"

Supplemental Figure 1. Mean cumulative function for the total number of skin cancers over time in the Medicare dataset.

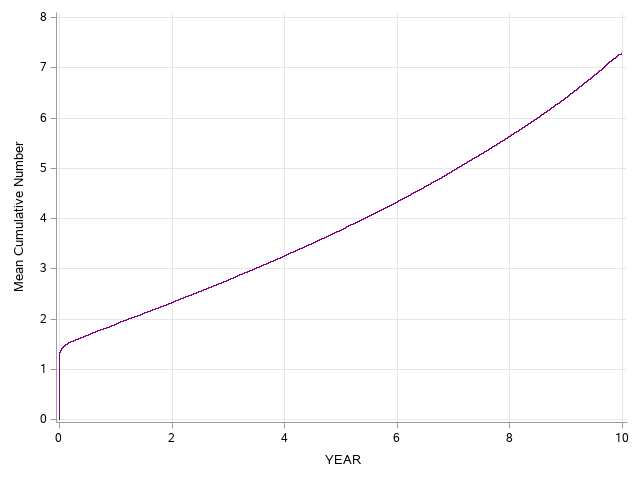

Supplemental Figure 2. Mean cumulative function for the total number of skin cancers over time in the Marketscan dataset.

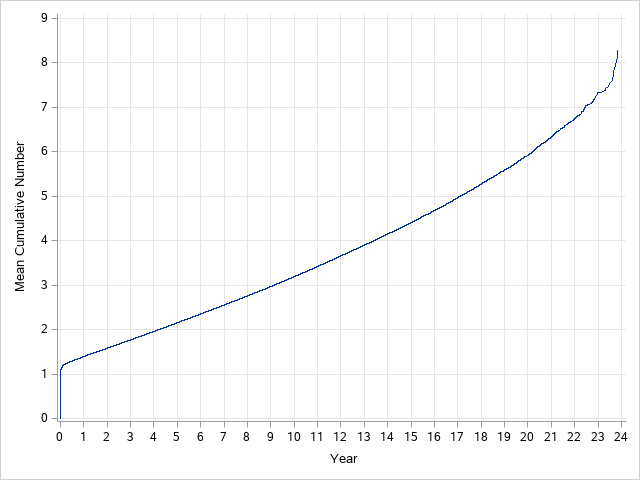

Supplemental Figure 3. Mean cumulative function for the total number of skin cancers over time in the Optum dataset.

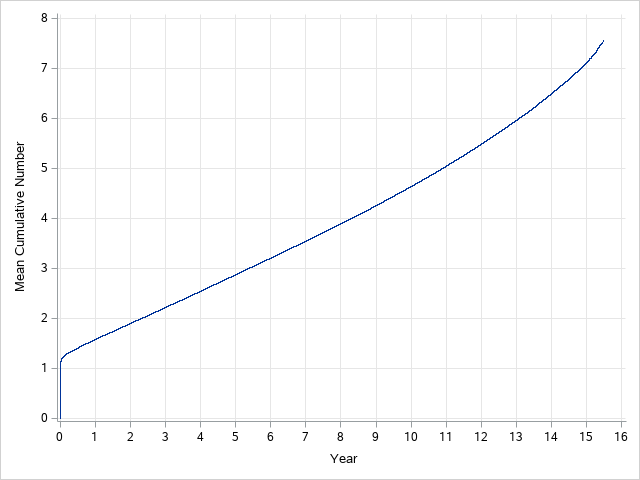

Supplemental Figure 4. Mean cumulative function for the total number of skin cancers over time in the Vanderbilt University Medical Center dataset.
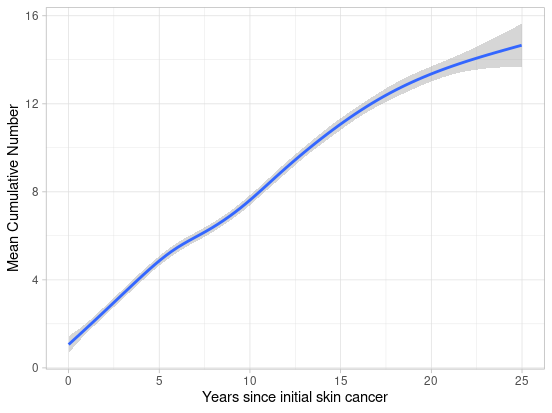

Supplemental Figure 5. Mean cumulative function for the total number of skin cancers over time in the VINCI dataset.

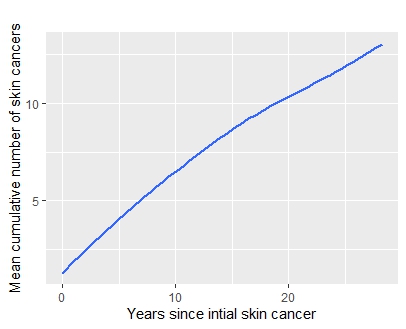

Supplemental Figure 6. Mean instantaneous rate of skin cancer treatment stratified by patients developing 2-3 skin cancers (red), 4-9 skin cancers (green), or 10 or more skin cancers (blue).

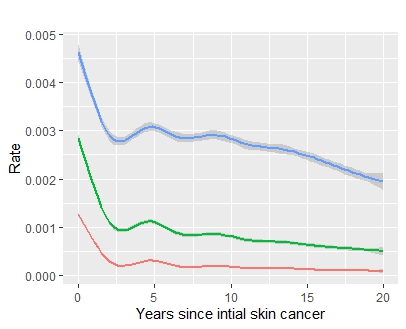
